# GLP-1 enhances beta-cell response to protein ingestion independent of glycemia and bariatric surgery amplifies it

**DOI:** 10.1101/2023.10.22.23297377

**Authors:** Maria Rayas, Amalia Gastaldelli, Henri Honka, Samantha Pezzica, Fabrizia Carli, Richard Peterson, Ralph DeFronzo, Marzieh Salehi

**Author notes:** Authors contributed equally and shared the first authorship position (Maria Rayas, Amalia Gastaldelli).

## Abstract

**Background:** The glycemic-independent actions of glucagon-like peptide 1 (GLP-1) in the prandial state in humans are largely unknown. Protein ingestion stimulates beta-cell secretion without changing plasma glucose concentration. We examined the contribution of endogenous GLP-1 to glucose metabolism and beta-cell response to protein ingestion under basal glucose concentrations, and whether these responses are affected by rerouted gut after gastric bypass (GB) or sleeve gastrectomy (SG).

**Methods:** Insulin secretion rate (ISR) and glucose fluxes during a 50-gram oral protein load were compared between 10 non-diabetic individuals with GB, 9 matched subjects with SG and 7 non-operated controls (CN) with and without intravenous infusion of exendin-(9–39) [Ex-9], a specific GLP-1 receptor (GLP-1R) antagonist.

**Results:** Blocking GLP-1R increased plasma glucose concentration before and after protein ingestion and decreased beta-cell sensitivity to glucose in the first 30 minutes of protein ingestion (p<0.05) in all 3 groups. However, reduction in the premeal ISR by Ex-9 infusion only was observed in CN (p<0.05 for interaction), whereas diminished prandial ISR_3h_ by GLP-1R blockade was observed in GB and SG and not in controls (p<0.05 for interaction). Also, GLP-1R blockade enhanced post-protein insulin action in GB and SG, but not in CN. Endogenous glucose production (*EGP*) during the first hour after protein ingestion was increased in all 3 groups but *EGP*_3h_ was accentuated by Ex-9 infusion only in GB (p<0.05 for interaction).

**Conclusion:** These findings are consistent with both a glucose-independent pancreatic and extra-pancreatic role for GLP-1 during protein ingestion in humans that are exaggerated by bariatric surgery.

**Trial registration:** This study was registered at Clinical Trials.Gov: NCT02823665

## INTRODUCTION

Glucagon-like peptide 1 (GLP-1), a proglucagon-derived peptide, plays a key role in normal glucose tolerance (1). GLP-1 is synthesized in L-cells of the intestinal mucosa, pancreatic alpha-cells, and neurons within the nucleus of the solitary tract in brain (1). After being released, GLP-1 acts through a specific GLP-1 receptor (GLP-1R) that is expressed in various tissues including islet cells and specific brain regions (1). Given that plasma GLP-1 concentration rises in proportion to the amount of ingested meal, it traditionally has been considered a hormone that communicates the information from the gut to the pancreatic islet-cells through the circulation.

The GLP-1-induced insulin secretion also has been presumed to be glucose dependent since exogenous infusion of GLP-1, which replicates prandial GLP-1 concentrations, has no insulinotropic effect under basal glucose concentrations (2, 3). Based on this conventional view, previous experiments, which used intravenous infusion of exendin-(9–39)[Ex-9], a potent GLP-1R antagonist, to demonstrate the insulin-stimulating and glucagon suppressive properties of *endogenous* GLP-1 in humans with (4) and without type 2 diabetes (T2D) (5, 6), all were conducted during a mixed meal or oral glucose load, where glycemia was above the baseline concentration.

Recently, the traditional concept that intestinally secreted GLP-1 acts on beta-cells in a glucose-dependent fashion to increase insulin secretion has been challenged. In healthy subjects and patients with T2D, blocking GLP-1R in the fasting state, when the plasma GLP-1 concentration is low, reduced basal insulin secretion (7–9). Further, blocking the GLP-1R during intravenous glucose infusion also diminished glucose-induced insulin secretion (4, 8, 10). These results suggest an important paracrine action of pancreatic produced GLP-1.

A case for paracrine or neural-mediated action of GLP-1 also has recently been implicated in the regulation of glucose tolerance in the fed state because of a small range of increase in prandial plasma GLP-1 concentration as well as rapid inactivation of GLP-1 in the circulation (11). However, the relevance of non-endocrine GLP-1 signals or glucose dependency of GLP-1 actions in prandial glucose metabolism in humans is unknown.

The hormonal versus non-hormonal actions of GLP-1 in the setting where meal-induced GLP-1 secretion is enhanced 5-10-fold, such as bariatric surgery, is also unexamined. The weight-loss independent glycemic effect of gastric bypass surgery (GB) and sleeve gastrectomy (SG) has been attributed, in part, to altered prandial nutrient flux/ metabolism, which is mediated by enhanced secretion of insulinotropic gut factors, mainly GLP-1(12–15). This conclusion is primarily based on previous reports that unanimously demonstrate that GLP-1R blockade during mixed meal or oral glucose ingestion has a greater effect to reduce insulin secretion in GB (13, 15–18) or SG (12), where both prandial GLP-1 secretion and glycemic excursion are augmented, compared to non-operated controls. However, recent preclinical studies have shown that pancreatic alpha-cell secretion of GLP-1, rather than intestinally produced peptide, plays a key role in beneficial glycemic effects of bariatric surgery (19). Consistent with these findings, in humans, despite a 5-to10-fold increase in intestinally derived GLP-1 secretion after GB or SG, there is no association between the magntidude of GLP-1-stimulated insulin secretion and the GLP-1 concentrations in prandial state (7). Further, in non-diabetic individuals, the beta-cell secretory response to increasing plasma concentrations of GLP-1 created by exogenous GLP-1 infusion (20) or mixed meal ingestion (21) is three-times smaller after GB compared to controls. Altogether, these observations suggest that non-hormonal actions of GLP-1 also play a role in altered glucose tolerance after GB or SG.

Finally, inhibition of prandial insulin secretion brought about by blocking GLP-1R after GB in subjects with or without T2D, is not associated with glycemic changes (13, 15–18). Therefore, it is plausible that GB elicits an extra-pancreatic action of GLP-1 (insulin action) that opposes the pancreatic effect of GLP-1 (insulin secretion).

The current studies therefore were undertaken to determine whether endogenous GLP-1 contributes to the insulinotropic effect of protein ingestion, where the plasma glucose concentration is maintained at euglycemic levels, and whether the prandial pancreatic (beta-cell) and extra-pancreatic (glucose flux) effects of endogenous GLP-1 are augmented after GB and SG given the enhanced GLP-1 secretion and nutrient flux following these procedures. To test these hypotheses, we examined the acute effect of GLP-1R blockade by administration of intravenous Ex-9 on glucose fluxes and islet-cell (insulin and glucagon) hormonal secretory responses to oral protein challenge in 3 groups of non-diabetic subjects: non-operated controls (CN), GB, and SG.

## RESULTS

### Subject characteristics (Table 1)

The GB, SG, and CN groups were similar in age, BMI, fat and lean mass, and female to male ratio. While the pre-operative BMI did not differ among GB and SG, % weight loss since surgery was larger in GB than SG, however, no significant differences were observed in pre-op BMI, weight loss and time post-surgery. HbA1c was lower in GB than SG and CN (p=0.05).

### Glucose concentrations

Baseline fasting and premeal glucose concentrations were similar among the groups (Table 2). Protein ingestion reduced prandial glucose values in controls by 0.1±0.05 mmol/l, but slightly raised the average plasma glucose concentration in GB and SG by 0.1±0.05 mmol/l, particularly in the first 60 minutes (Fig.1a; p<0.05).

**Table 1.**
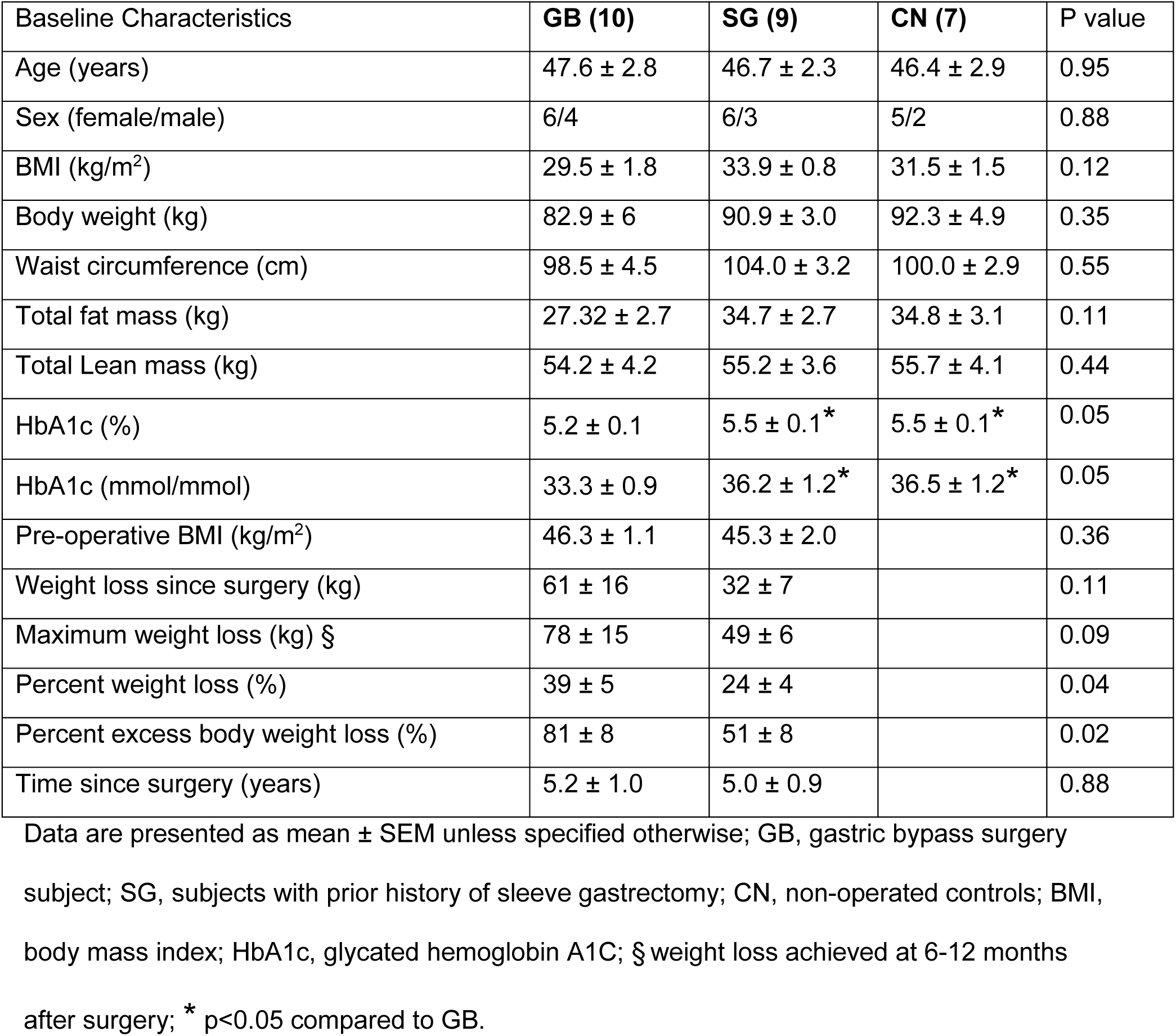
Baseline characteristics of study subjects.

| Baseline Characteristics | GB (10) | SG (9) | CN (7) | P value |
| --- | --- | --- | --- | --- |
| Age (years) | 47.6 ± 2.8 | 46.7 ± 2.3 | 46.4 ± 2.9 | 0.95 |
| Sex (female/male) | 6/4 | 6/3 | 5/2 | 0.88 |
| BMI (kg/m<sup>2</sup>) | 29.5 ± 1.8 | 33.9 ± 0.8 | 31.5 ± 1.5 | 0.12 |
| Body weight (kg) | 82.9 ± 6 | 90.9 ± 3.0 | 92.3 ± 4.9 | 0.35 |
| Waist circumference (cm) | 98.5 ± 4.5 | 104.0 ± 3.2 | 100.0 ± 2.9 | 0.55 |
| Total fat mass (kg) | 27.32 ± 2.7 | 34.7 ± 2.7 | 34.8 ± 3.1 | 0.11 |
| Total Lean mass (kg) | 54.2 ± 4.2 | 55.2 ± 3.6 | 55.7 ± 4.1 | 0.44 |
| HbA1c (%) | 5.2 ± 0.1 | 5.5 ± 0.1<sup>*</sup> | 5.5 ± 0.1<sup>*</sup> | 0.05 |
| HbA1c (mmol/mmol) | 33.3 ± 0.9 | 36.2 ± 1.2<sup>*</sup> | 36.5 ± 1.2<sup>*</sup> | 0.05 |
| Pre-operative BMI (kg/m<sup>2</sup>) | 46.3 ± 1.1 | 45.3 ± 2.0 |  | 0.36 |
| Weight loss since surgery (kg) | 61 ± 16 | 32 ± 7 |  | 0.11 |
| Maximum weight loss (kg) § | 78 ± 15 | 49 ± 6 |  | 0.09 |
| Percent weight loss (%) | 39 ± 5 | 24 ± 4 |  | 0.04 |
| Percent excess body weight loss (%) | 81 ± 8 | 51 ± 8 |  | 0.02 |
| Time since surgery (years) | 5.2 ± 1.0 | 5.0 ± 0.9 |  | 0.88 |

Data are presented as mean ± SEM unless specified otherwise; GB, gastric bypass surgery subject; SG, subjects with prior history of sleeve gastrectomy; CN, non-operated controls; BMI, body mass index; HbA1c, glycated hemoglobin A1C; § weight loss achieved at 6-12 months after surgery; \* p<0.05 compared to GB.

**Table 2.**
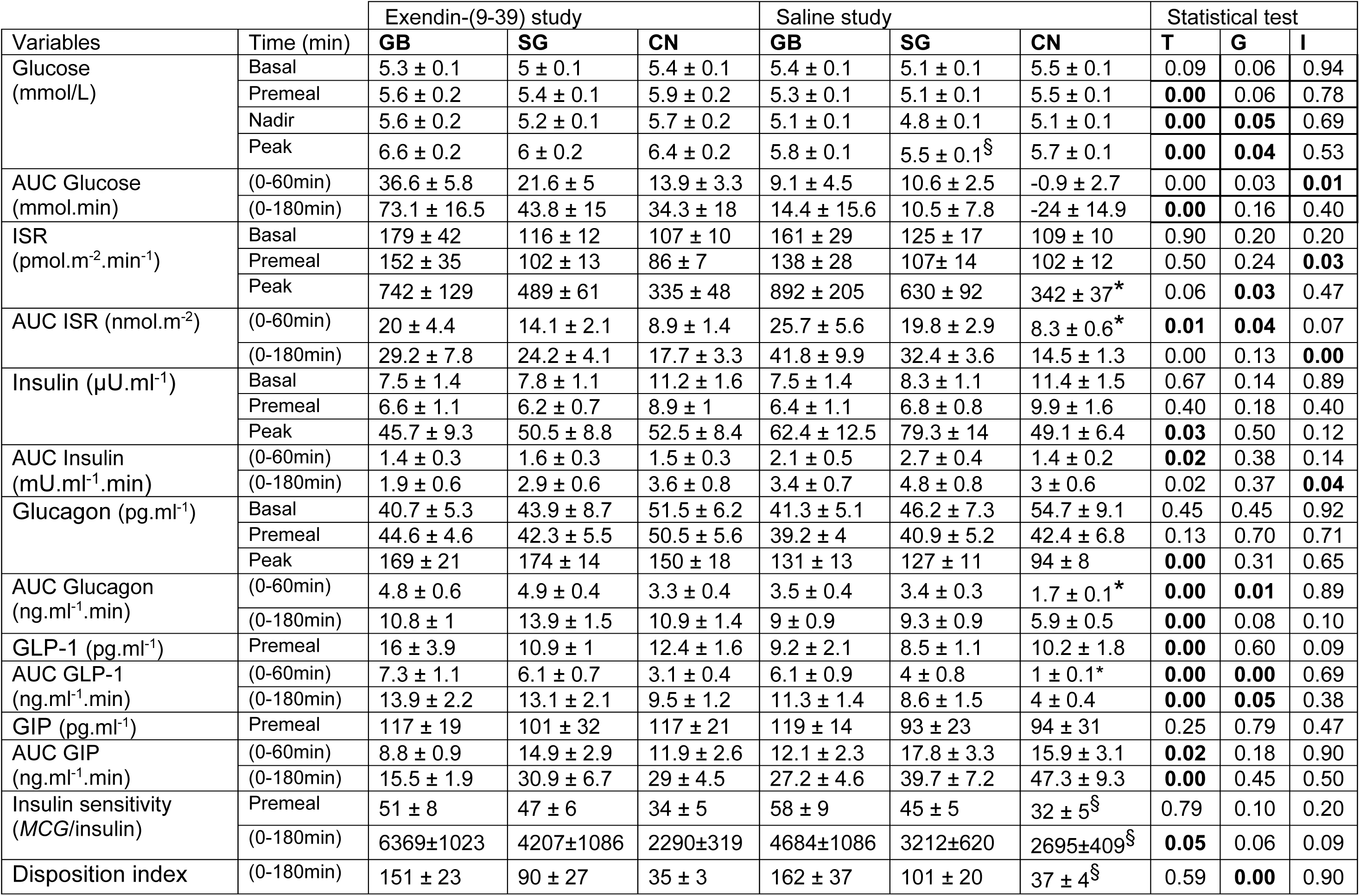
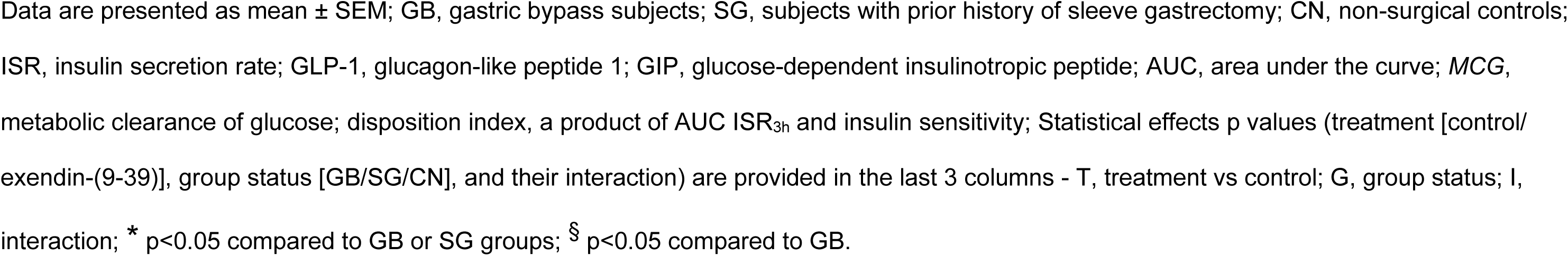
Glucose, islet-cell, and incretin secretory responses to protein ingestion with and without GLP-1R blockade in GB, SG and CN subjects.

| Variables | Time (min) | Exendin-(9-39) study |  |  | Saline study |  |  | Statistical test |  |  |
| --- | --- | --- | --- | --- | --- | --- | --- | --- | --- | --- |
|  |  | GB | SG | CN | GB | SG | CN | T | G | I |
| Glucose (mmol/L) | Basal | 5.3 ± 0.1 | 5 ± 0.1 | 5.4 ± 0.1 | 5.4 ± 0.1 | 5.1 ± 0.1 | 5.5 ± 0.1 | 0.09 | 0.06 | 0.94 |
|  | Premeal | 5.6 ± 0.2 | 5.4 ± 0.1 | 5.9 ± 0.2 | 5.3 ± 0.1 | 5.1 ± 0.1 | 5.5 ± 0.1 | **0.00** | 0.06 | 0.78 |
|  | Nadir | 5.6 ± 0.2 | 5.2 ± 0.1 | 5.7 ± 0.2 | 5.1 ± 0.1 | 4.8 ± 0.1 | 5.1 ± 0.1 | **0.00** | **0.05** | 0.69 |
|  | Peak | 6.6 ± 0.2 | 6 ± 0.2 | 6.4 ± 0.2 | 5.8 ± 0.1 | 5.5 ± 0.1<sup>§</sup> | 5.7 ± 0.1 | **0.00** | **0.04** | 0.53 |
| AUC Glucose (mmol.min) | (0-60min) | 36.6 ± 5.8 | 21.6 ± 5 | 13.9 ± 3.3 | 9.1 ± 4.5 | 10.6 ± 2.5 | -0.9 ± 2.7 | 0.00 | 0.03 | **0.01** |
|  | (0-180min) | 73.1 ± 16.5 | 43.8 ± 15 | 34.3 ± 18 | 14.4 ± 15.6 | 10.5 ± 7.8 | -24 ± 14.9 | **0.00** | 0.16 | 0.40 |
| ISR (pmol.m<sup>-2</sup>.min<sup>-1</sup>) | Basal | 179 ± 42 | 116 ± 12 | 107 ± 10 | 161 ± 29 | 125 ± 17 | 109 ± 10 | 0.90 | 0.20 | 0.20 |
|  | Premeal | 152 ± 35 | 102 ± 13 | 86 ± 7 | 138 ± 28 | 107 ± 14 | 102 ± 12 | 0.50 | 0.24 | **0.03** |
|  | Peak | 742 ± 129 | 489 ± 61 | 335 ± 48 | 892 ± 205 | 630 ± 92 | 342 ± 37* | 0.06 | **0.03** | 0.47 |
| AUC ISR (nmol.m<sup>-2</sup>) | (0-60min) | 20 ± 4.4 | 14.1 ± 2.1 | 8.9 ± 1.4 | 25.7 ± 5.6 | 19.8 ± 2.9 | 8.3 ± 0.6* | **0.01** | **0.04** | 0.07 |
|  | (0-180min) | 29.2 ± 7.8 | 24.2 ± 4.1 | 17.7 ± 3.3 | 41.8 ± 9.9 | 32.4 ± 3.6 | 14.5 ± 1.3 | 0.00 | 0.13 | **0.00** |
| Insulin (μU.ml<sup>-1</sup>) | Basal | 7.5 ± 1.4 | 7.8 ± 1.1 | 11.2 ± 1.6 | 7.5 ± 1.4 | 8.3 ± 1.1 | 11.4 ± 1.5 | 0.67 | 0.14 | 0.89 |
|  | Premeal | 6.6 ± 1.1 | 6.2 ± 0.7 | 8.9 ± 1 | 6.4 ± 1.1 | 6.8 ± 0.8 | 9.9 ± 1.6 | 0.40 | 0.18 | 0.40 |
|  | Peak | 45.7 ± 9.3 | 50.5 ± 8.8 | 52.5 ± 8.4 | 62.4 ± 12.5 | 79.3 ± 14 | 49.1 ± 6.4 | **0.03** | 0.50 | 0.12 |
| AUC Insulin (mU.ml<sup>-1</sup>.min) | (0-60min) | 1.4 ± 0.3 | 1.6 ± 0.3 | 1.5 ± 0.3 | 2.1 ± 0.5 | 2.7 ± 0.4 | 1.4 ± 0.2 | **0.02** | 0.38 | 0.14 |
|  | (0-180min) | 1.9 ± 0.6 | 2.9 ± 0.6 | 3.6 ± 0.8 | 3.4 ± 0.7 | 4.8 ± 0.8 | 3 ± 0.6 | 0.02 | 0.37 | **0.04** |
| Glucagon (pg.ml<sup>-1</sup>) | Basal | 40.7 ± 5.3 | 43.9 ± 8.7 | 51.5 ± 6.2 | 41.3 ± 5.1 | 46.2 ± 7.3 | 54.7 ± 9.1 | 0.45 | 0.45 | 0.92 |
|  | Premeal | 44.6 ± 4.6 | 42.3 ± 5.5 | 50.5 ± 5.6 | 39.2 ± 4 | 40.9 ± 5.2 | 42.4 ± 6.8 | 0.13 | 0.70 | 0.71 |
|  | Peak | 169 ± 21 | 174 ± 14 | 150 ± 18 | 131 ± 13 | 127 ± 11 | 94 ± 8 | **0.00** | 0.31 | 0.65 |
| AUC Glucagon (ng.ml<sup>-1</sup>.min) | (0-60min) | 4.8 ± 0.6 | 4.9 ± 0.4 | 3.3 ± 0.4 | 3.5 ± 0.4 | 3.4 ± 0.3 | 1.7 ± 0.1* | **0.00** | **0.01** | 0.89 |
|  | (0-180min) | 10.8 ± 1 | 13.9 ± 1.5 | 10.9 ± 1.4 | 9 ± 0.9 | 9.3 ± 0.9 | 5.9 ± 0.5 | **0.00** | 0.08 | 0.10 |
| GLP-1 (pg.ml<sup>-1</sup>) | Premeal | 16 ± 3.9 | 10.9 ± 1 | 12.4 ± 1.6 | 9.2 ± 2.1 | 8.5 ± 1.1 | 10.2 ± 1.8 | **0.00** | 0.60 | 0.09 |
| AUC GLP-1 (ng.ml<sup>-1</sup>.min) | (0-60min) | 7.3 ± 1.1 | 6.1 ± 0.7 | 3.1 ± 0.4 | 6.1 ± 0.9 | 4 ± 0.8 | 1 ± 0.1* | **0.00** | **0.00** | 0.69 |
|  | (0-180min) | 13.9 ± 2.2 | 13.1 ± 2.1 | 9.5 ± 1.2 | 11.3 ± 1.4 | 8.6 ± 1.5 | 4 ± 0.4 | **0.00** | **0.05** | 0.38 |
| GIP (pg.ml<sup>-1</sup>) | Premeal | 117 ± 19 | 101 ± 32 | 117 ± 21 | 119 ± 14 | 93 ± 23 | 94 ± 31 | 0.25 | 0.79 | 0.47 |
| AUC GIP (ng.ml<sup>-1</sup>.min) | (0-60min) | 8.8 ± 0.9 | 14.9 ± 2.9 | 11.9 ± 2.6 | 12.1 ± 2.3 | 17.8 ± 3.3 | 15.9 ± 3.1 | **0.02** | 0.18 | 0.90 |
|  | (0-180min) | 15.5 ± 1.9 | 30.9 ± 6.7 | 29 ± 4.5 | 27.2 ± 4.6 | 39.7 ± 7.2 | 47.3 ± 9.3 | **0.00** | 0.45 | 0.50 |
| Insulin sensitivity (MCG/insulin) | Premeal | 51 ± 8 | 47 ± 6 | 34 ± 5 | 58 ± 9 | 45 ± 5 | 32 ± 5<sup>§</sup> | 0.79 | 0.10 | 0.20 |
|  | (0-180min) | 6369±1023 | 4207±1086 | 2290±319 | 4684±1086 | 3212±620 | 2695±409<sup>§</sup> | **0.05** | 0.06 | 0.09 |
| Disposition index | (0-180min) | 151 ± 23 | 90 ± 27 | 35 ± 3 | 162 ± 37 | 101 ± 20 | 37 ± 4<sup>§</sup> | 0.59 | **0.00** | 0.90 |

Data are presented as mean ± SEM; GB, gastric bypass subjects; SG, subjects with prior history of sleeve gastrectomy; CN, non-surgical controls; ISR, insulin secretion rate; GLP-1, glucagon-like peptide 1; GIP, glucose-dependent insulintropic peptide; AUC, area under the curve; *MCG*, metabolic clearance of glucose; disposition index, a product of AUC ISR<sub>3h</sub> and insulin sensitivity; Statistical effects p values (treatment [control/exendin-(9-39)], group status [GB/SG/CN], and their interaction) are provided in the last 3 columns - T, treatment vs control; G, group status; I, interaction; \* p<0.05 compared to GB or SG groups; § p<0.05 compared to GB.

Blocking GLP-1R similarly increased average plasma glucose concentrations before and after protein ingestion by 5-6% and 10-15%, respectively, in all 3 groups (Fig.1a; p<0.05). However, the early glycemic effect of Ex-9 after protein intake (AUC Glucose_1h_) was much more robust in GB and SG compared to non-operated controls (Table 2; p<0.05 for interaction).

### Beta- and alpha-cell responses

Baseline fasting levels of insulin and ISR were similar among the 3 groups and between the two studies (Table 2). During saline studies, protein ingestion increased beta-cell secretion (AUC ISR_3h_) in all 3 groups, but due to a shift of ISR response to the left, the AUC ISR_1h_ was larger in surgical compared to controls (Table 2, Fig.1b; p<0.05). Also, disposition index (DI) calculated as product of AUC ISR_3h_ and insulin sensitivity was significantly larger in GB versus SG or CN (Table 2; p<0.05).

**Figure 1.**
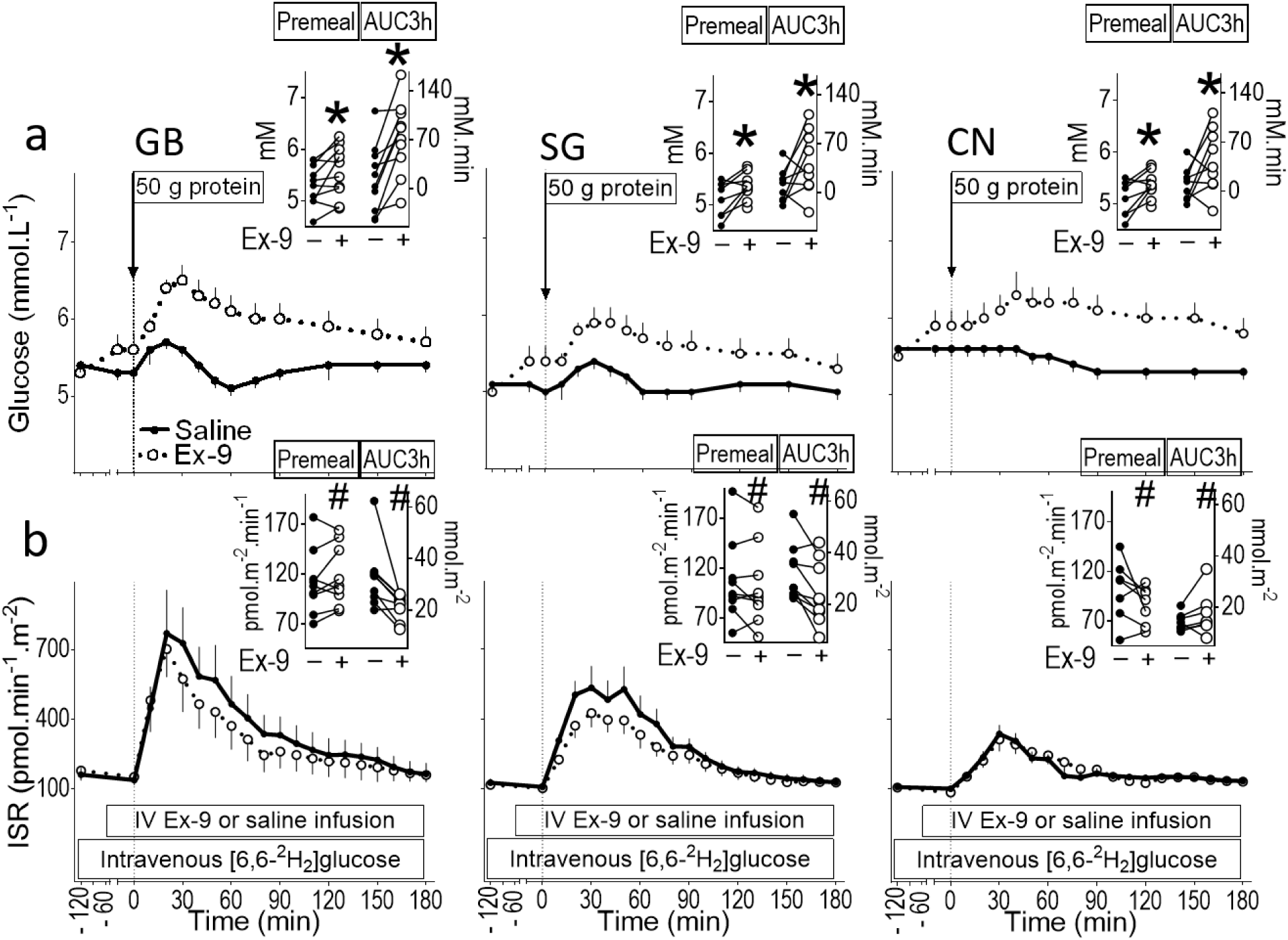
(a) Plasma glucose concentration and (b) insulin secretion rate (ISR) before and after oral protein ingestion with intravenous infusion of saline (solid line) or exendin-(9–39) (Ex-9) (dashed line), in subjects who underwent gastric bypass (left panel) or sleeve gastrectomy (middle panel) and non-operated controls (right panel). The corresponding individual changes from saline to Ex-9 study for premeal and AUC 3h values are shown (insets). *P < .05 compared with saline study; # P < 0.05 for interaction.

**Figure 2.**
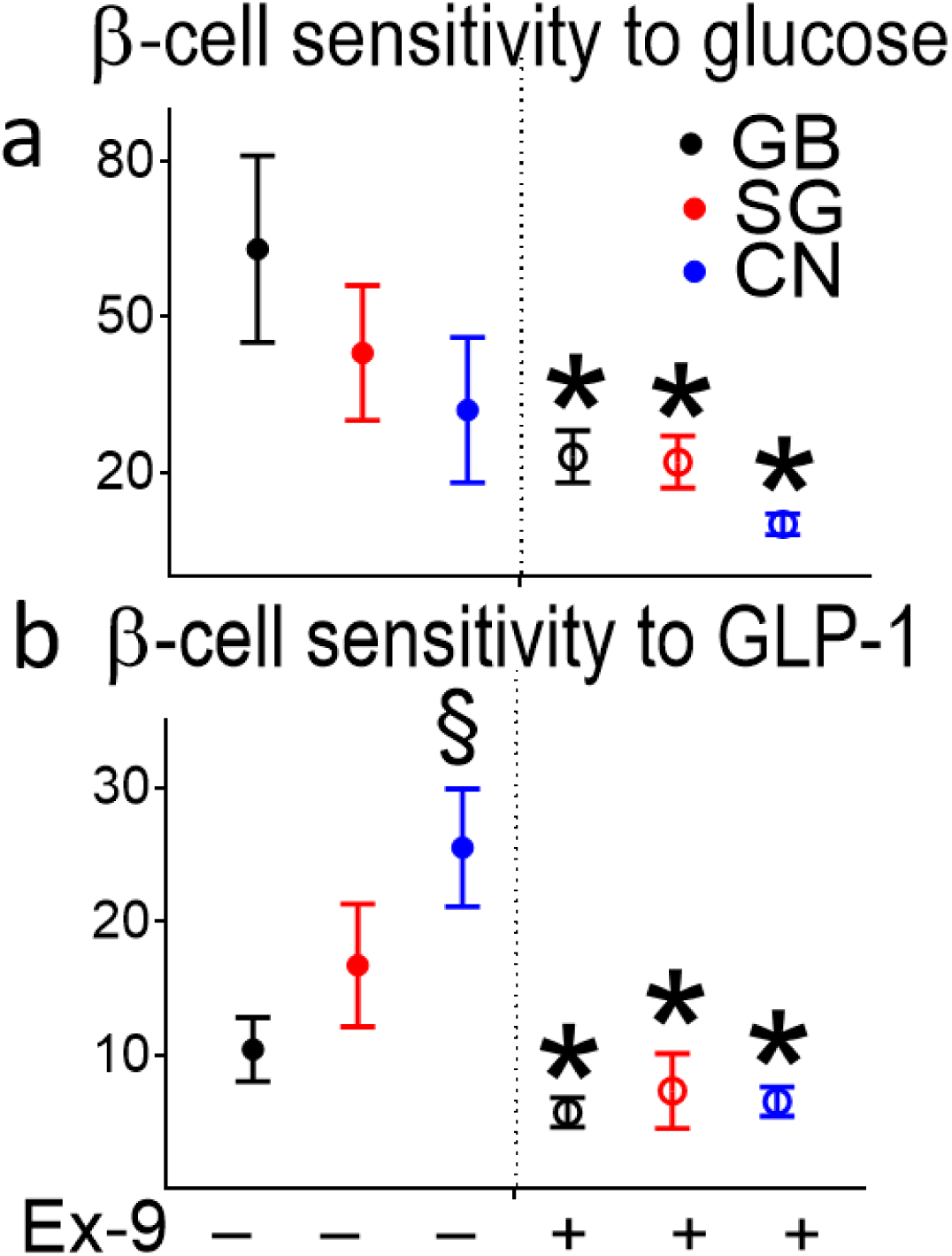
Beta-cell sensitivity to (a) glucose and (b) GLP-1 measured as the slope of ISR (from premeal to peak values) versus corresponding plasma concentration of glucose and GLP-1, respectively during saline (left panel) and GLP-1R blockade (Ex-9) (right panel) conditions in subjects who underwent gastric bypass (black line) or sleeve gastrectomy (red line) and non-operated controls (blue line). * P < 0.05 compared with saline study; § P< 0.05 compared with GB; # P < 0.05 for interaction.

Blocking GLP-1R tended to decrease premeal ISR levels in CN despite an increase in glycemia but not in GB or SG subjects (Table 2; p<0.05 for interaction); the relative change in premeal ISR from saline to Ex-9 studies in GB and SG versus CN was 9±4 % and -4±6 % versus -11±8

% (Fig.1b inset). In contrast, postprandial beta-cell secretory response was reduced only in surgical subjects by Ex-9 infusion (Table 2; p<0.05 for interaction); the relative change in AUC ISR_3h_ from saline to Ex-9 studies in GB and SG versus CN was - 29±6% and - 27±8% versus 19±13% (Fig.1b inset).

Beta-cell glucose sensitivity during the first part of protein absorption, where ISR rose from premeal to peak value, however, did not differ among surgical and non-surgical controls and similarly diminished in all 3 groups by blocking GLP-1R (Fig.2a; p<0.05).

Beta-cell responsiveness to increasing plasma concentrations of GLP-1 from premeal to peak value during protein ingestion was significantly larger in CN versus GB during saline study (Fig.2b; p<0.05). As expected, blocking GLP-1R markedly decreased the ISR response to increasing plasma GLP-1 concentrations in all 3 groups (Fig.2b; p<0.01).

Baseline and premeal glucagon levels were similar among the 3 groups and between the two studies of Ex-9 or saline infusion (Table 2). During saline study, early glucagon response to protein ingestion (AUC Glucagon_1h_) was larger in surgical than CN (Fig.3a; p<0.05) with a similar trend noted over 180 minutes (AUC Glucagon_3h_) (Fig.3a; p=0.08). Ex-9 infusion similarly increased plasma concentrations of glucagon after protein ingestion in all 3 groups (Table 2, Fig.3a; p<0.05).

**Figure 3.**
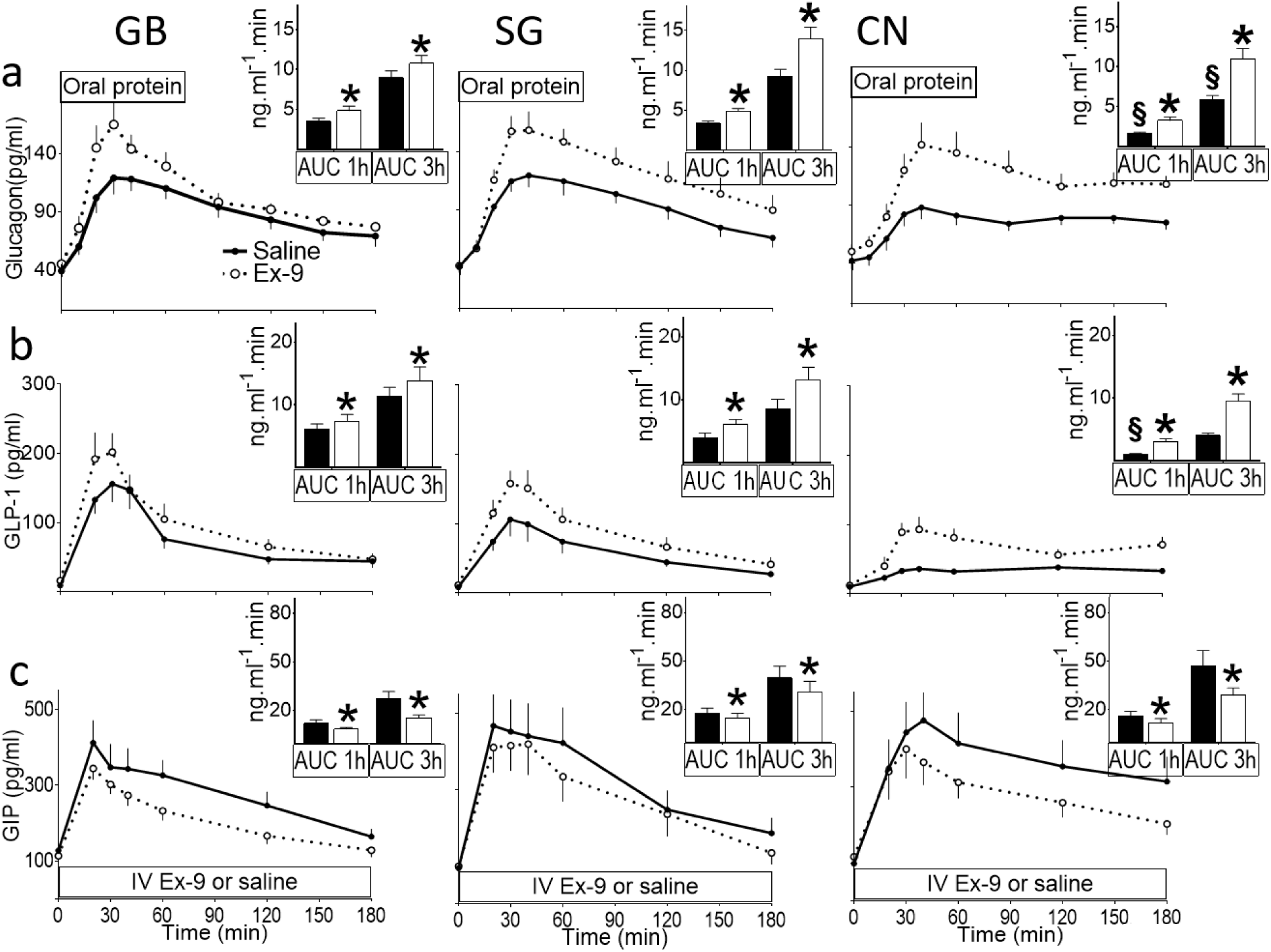
Plasma concentrations of (a) glucagon, (b) GLP-1, and (c) GIP during protein ingestion with (dashed line) and without (solid line) intravenous infusion of exendin-(9–39) (Ex-9), in subjects who underwent gastric bypass (left panel) or sleeve gastrectomy (middle panel) and non-operated controls (right panel). The corresponding AUCs from 0 to 60 min and from 0 to 180 min are shown (insets). * P < 0.05 compared with saline study; § P< 0.05 compared with GB or SG.

During saline studies, the plasma glucagon-to-insulin ratio showed an early reduction within the first 30-60 min in all 3 groups with a reversal to premeal values by 3 hours from protein intake, although higher in GB (Supplementary.Fig1a). Blocking GLP-1 had no effect on the plasma glucagon-to-insulin ratio in healthy controls, but significantly raised this ratio beyond the first 30 min of protein ingestion in both GB and SG (Supplementary.Fig1), likely due to a larger shift in beta-cell secretory response.

### Incretin response

Fasting levels of GLP-1 and GIP were similar among 3 groups. Ex-9 infusion increased premeal concentrations of GLP-1 but not GIP (Table 2).

Protein consumption raised plasma GLP-1 concentrations in all 3 groups, but to much larger extent in GB and SG than CN (Fig.3b; p<0.05); GLP-1R blockade further increased GLP-1 response to protein intake, which was much greater in CN than SG or GB (relative increase in AUC3h: 30±17, 64±22, and 141±26 % in GB, SG, and CN; p<0.001). The GIP secretory effect of protein ingestion did not differ across the groups (Table 2; Fig.3c) and Ex-9 infusion decreased GIP secretion in all 3 groups (Fig.3c; p<0.05). The magnitude of reduction in AUC ISR_3h_ by Ex-9 infusion did not correlate with the size of increased plasma GLP-1 concentrations during control or Ex-9 studies.

### Gastric emptying

Time to peak plasma ingested acetaminophen concentration was shorter in GB and SG compared to CN (34±11, 56±21, and 148±17 min in GB, SG, and CN; p<0.001) and C_max_ was larger in surgical than controls (103±14, 76±9, and 43±4 µmol/L in GB, SG, and CN; p<0.001), but neither was affected by Ex-9 infusion (Supplementary.Fig2).

### Glucose kinetics

Following an overnight fast, under the steady-state condition, the rate of total body glucose utilization (*Rd*) equals the rate of endogenous glucose production (*EGP*), and was similar among GB, SG, and controls (Table 3; Fig.4).

**Figure 4.**
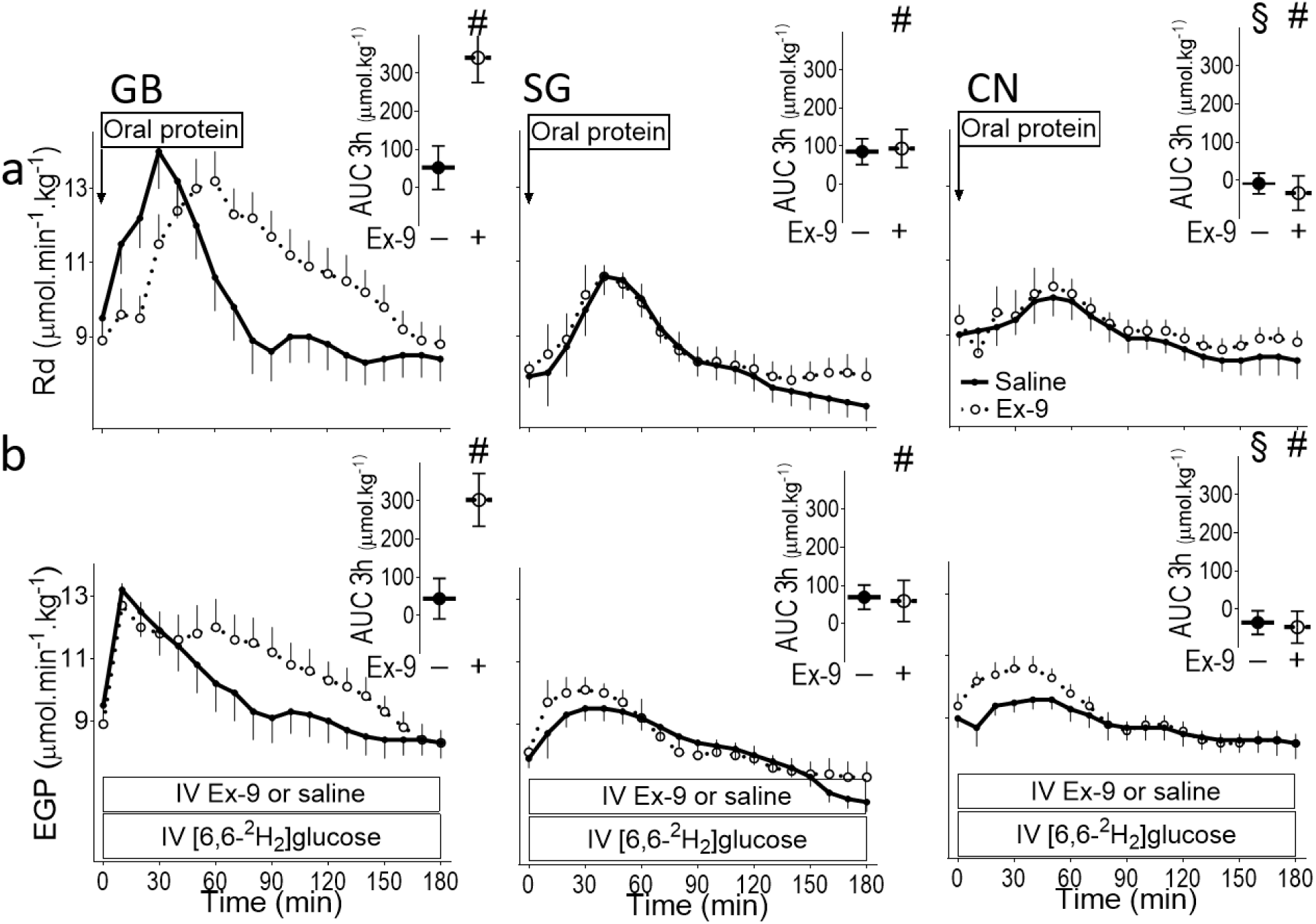
The rates of (a) total glucose utilization (*Rd*), (b) endogenous glucose production (*EGP*) during oral protein ingestion with (dashed line) and without (solid line) intravenous infusion of exendin-(9–39) (Ex-9) in subjects who underwent gastric bypass (left panel) or sleeve gastrectomy (middle panel) and non-operated controls (right panel). The corresponding AUCs from 0 to 180 min are shown (insets). § P< 0.05 compared with GB or SG; # P < 0.05 for interaction.

**Table 3.**
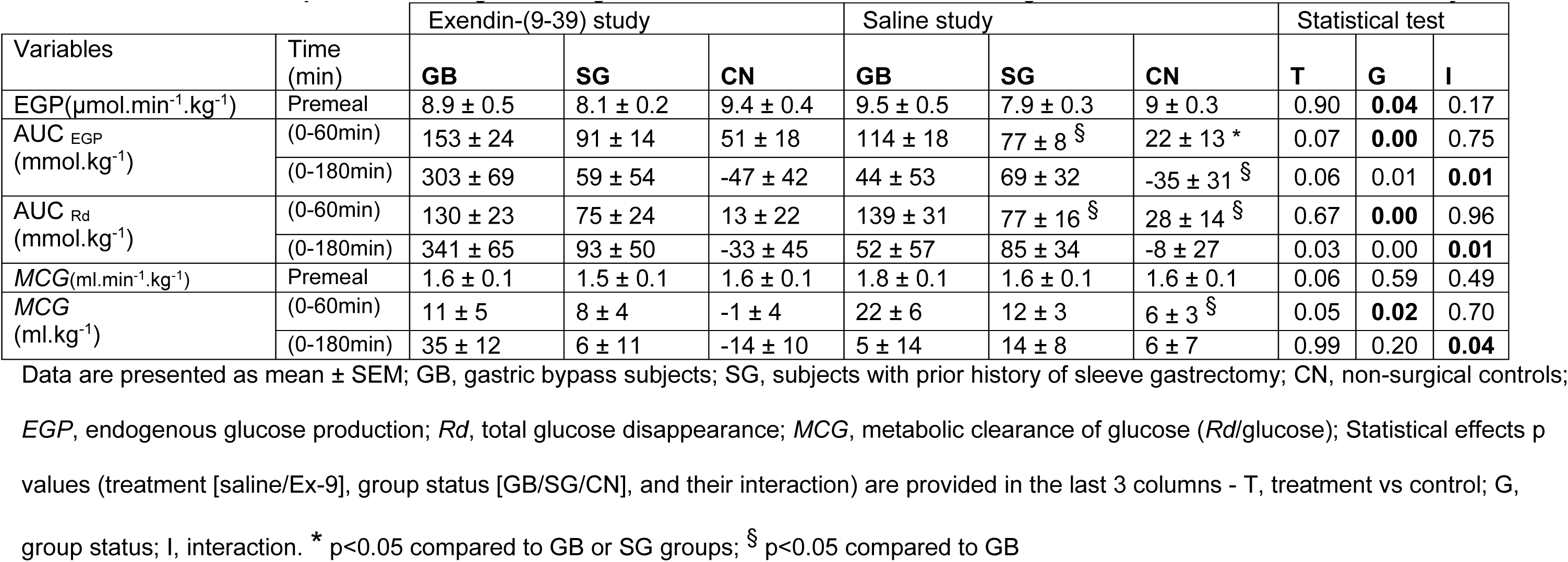
Glucose flux responses to oral glucose ingestion with and without GLP-1R antagonist infusion in GB, SG and CN subjects.

| Variables | Time (min) | Exendin-(9-39) study |  |  | Saline study |  |  | Statistical test |  |  |
| --- | --- | --- | --- | --- | --- | --- | --- | --- | --- | --- |
|  |  | GB | SG | CN | GB | SG | CN | T | G | I |
| EGP($\mu\text{mol}\cdot\text{min}^{-1}\cdot\text{kg}^{-1}$) | Premeal | 8.9 ± 0.5 | 8.1 ± 0.2 | 9.4 ± 0.4 | 9.5 ± 0.5 | 7.9 ± 0.3 | 9 ± 0.3 | 0.90 | **0.04** | 0.17 |
| AUC<sub>EGP</sub><br>($\text{mmol}\cdot\text{kg}^{-1}$) | (0-60min) | 153 ± 24 | 91 ± 14 | 51 ± 18 | 114 ± 18 | 77 ± 8<sup>§</sup> | 22 ± 13 * | 0.07 | **0.00** | 0.75 |
| | (0-180min) | 303 ± 69 | 59 ± 54 | -47 ± 42 | 44 ± 53 | 69 ± 32 | -35 ± 31<sup>§</sup> | 0.06 | 0.01 | **0.01** |
| AUC<sub>Rd</sub><br>($\text{mmol}\cdot\text{kg}^{-1}$) | (0-60min) | 130 ± 23 | 75 ± 24 | 13 ± 22 | 139 ± 31 | 77 ± 16<sup>§</sup> | 28 ± 14<sup>§</sup> | 0.67 | **0.00** | 0.96 |
| | (0-180min) | 341 ± 65 | 93 ± 50 | -33 ± 45 | 52 ± 57 | 85 ± 34 | -8 ± 27 | 0.03 | 0.00 | **0.01** |
| MCG($\text{ml}\cdot\text{min}^{-1}\cdot\text{kg}^{-1}$) | Premeal | 1.6 ± 0.1 | 1.5 ± 0.1 | 1.6 ± 0.1 | 1.8 ± 0.1 | 1.6 ± 0.1 | 1.6 ± 0.1 | 0.06 | 0.59 | 0.49 |
| MCG<br>($\text{ml}\cdot\text{kg}^{-1}$) | (0-60min) | 11 ± 5 | 8 ± 4 | -1 ± 4 | 22 ± 6 | 12 ± 3 | 6 ± 3<sup>§</sup> | 0.05 | **0.02** | 0.70 |
| | (0-180min) | 35 ± 12 | 6 ± 11 | -14 ± 10 | 5 ± 14 | 14 ± 8 | 6 ± 7 | 0.99 | 0.20 | **0.04** |

Data are presented as mean ± SEM; GB, gastric bypass subjects; SG, subjects with prior history of sleeve gastrectomy; CN, non-surgical controls;
EGP, endogenous glucose production; Rd, total glucose disappearance; MCG, metabolic clearance of glucose (Rd/glucose); Statistical effects p
values (treatment [saline/Ex-9], group status [GB/SG/CN], and their interaction) are provided in the last 3 columns - T, treatment vs control; G,
group status; I, interaction. \* p<0.05 compared to GB or SG groups; § p<0.05 compared to GB

In response to protein ingestion, *EGP* rose but to a much larger extent in GB and SG than CN (Table 3, Fig.4b; p<0.05). Ex-9 infusion tended to increase the early prandial *EGP* (AUC *EGP*_1h_) in all 3 groups (Table 3; p=0.07). However, the overall *EGP* following oral protein (AUC *EGP*_3h_) during Ex-9 studies was increased only in GB and not in SG or CN (Table 3, Fig.4b; p<0.05 for interaction).

Following protein ingestion, in parallel with *EGP* response, incremental rates of glucose disposal (*Rd*) were larger in surgical, especially GB, than in CN (Table3, Fig.4a; p<0.05). However, metabolic clearance of glucose (*MCG*), i.e., *Rd* adjusted for glucose levels, over the 3 hours from protein intake was not significantly different among 3 groups (Table 3). Blocking GLP-1R augmented AUC *Rd_3h_* or AUC *MCG_3h_* only in GB subjects without any significant effect in SG or CN (Table 3, Fig.4a; p<0.01 for interaction).

### Insulin action

Before and after protein ingestion whole body insulin action on glucose metabolism, measured by premeal *MCG*/insulin and AUC *MCG*/insulin_3h_, respectively, were greater in GB compared to controls (Table 2, Fig.5b; p<0.05). GLP-1R antagonist had no effect on premeal insulin action but increased prandial TAUC *MCG*/insulin_3h_ in surgical, particularly in GB subjects compared to controls (Table 2, Fig.5b; p<0.05).

**Figure 5.**
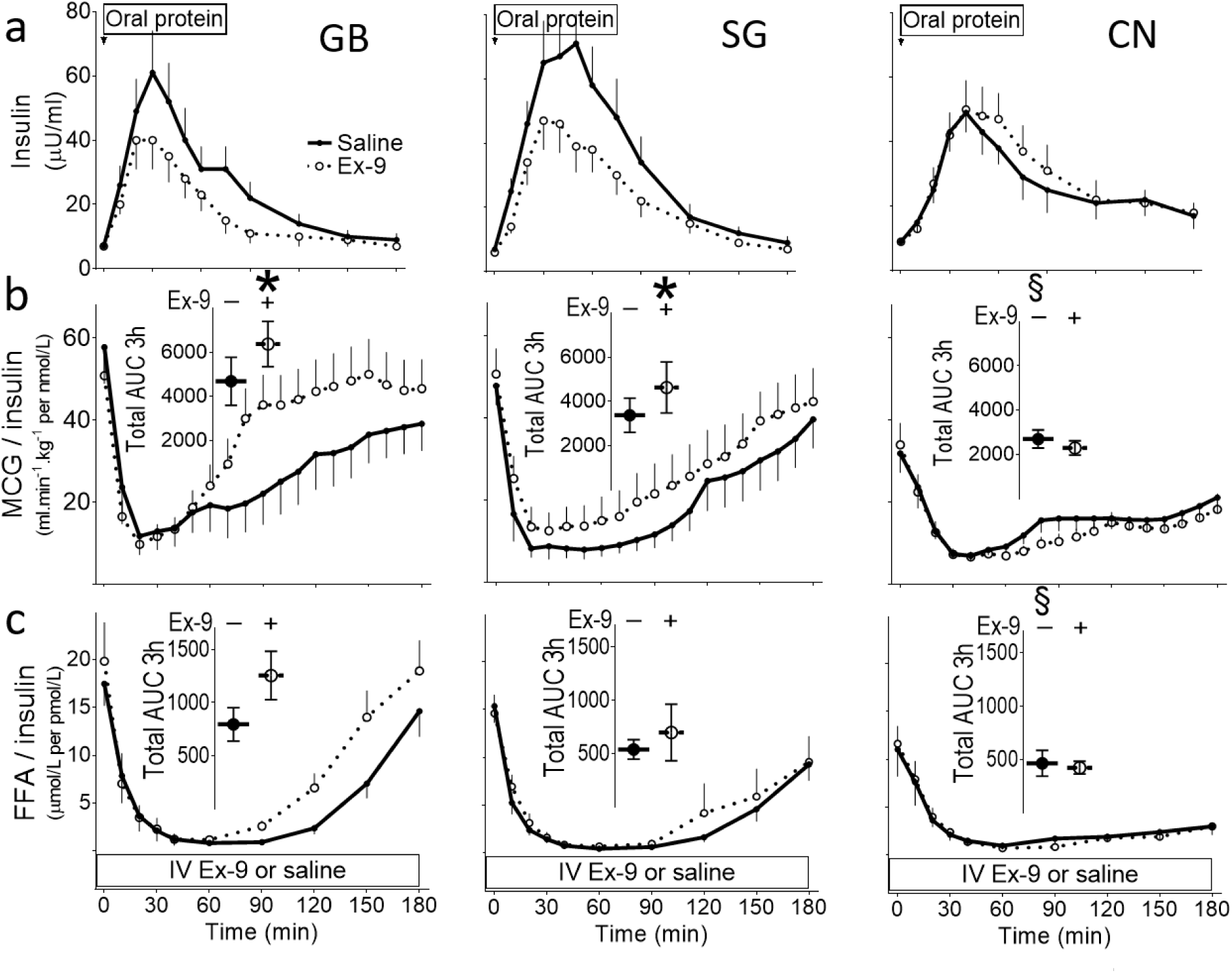
(a) Plasma concentrations of insulin, (b) rates of metabolic glucose clearance (MCG) adjusted for insulin concentration, and (c) plasma free fatty acid concentrations (FFA) adjusted for insulin levels with (dashed line) and without (solid line) GLP-1R blockade (Ex-9), in subjects who underwent gastric bypass (left panel) or sleeve gastrectomy (middle panel) and non-operated controls (right panel). The corresponding prandial AUCs from 0 to 180 min are shown (insets). * P < 0.05 compared with saline study; § P< 0.05 compared with GB.

Insulin action in suppressing FFA at baseline, measured by premeal FFA/insulin, did not differ among 3 groups (Fig.5c). Prandial AUC FFA/insulin_3h_, however, tended to be larger in GB than SG or CN (p=0.06) and increased further by Ex-9 infusion in GB subjects compared to GB or CN (Fig.5c; p=0.07 for interaction).

## DISCUSSION

The findings reported here demonstrate a novel insulinotropic effect of endogenous GLP-1 during protein ingestion in humans, where glucose concentration is maintained at basal level. Also, we have shown that the rerouted GI anatomy after GB and SG enhances GLP-1-stimulated beta-cell response to protein ingestion. Importantly, in our experiment, beta cell sensitivity to GLP-1 (Fig.2b) and to glucose (Fig.2a) following protein ingestion were markedly reduced by Ex-9, demonstrating a causative role for GLP-1. However, there was no relationship between the magnitude of the GLP-1 effect on insulin secretion and plasma GLP-1 concentrations, which mainly reflects intestinally produced peptide. Lastly, we also have observed that GLP-1R blockade increased insulin action in skeletal muscle in subjects with prior history of bariatric surgery, particularly after gastric bypass (Fig.5b). Together, these observations in the context of recent clinical (9, 22) and preclinical studies(19, 23): (*1*) highlight the significance of GLP-1R signal in regulation of glucose metabolism during both fasting and prandial state independent of plasma glucose or GLP-1 concentrations, (*2*) are consistent with a model of non-endocrine effect of GLP-1 mediated by either CNS regulation of glucose-stimulated insulin secretion and glucose flux (more relevant to bariatric subjects) or paracrine regulation of beta-cell response (more relevant to non-operated controls) or both in response to acute stimulus of orally ingested protein, and (*3*) indicate that rerouted gut after bariatric surgery, particularly gastric bypass, alters both pancreatic and extra-pancreatic GLP-1 action during protein intake.

Using Ex-9, we (4, 6) and others (5) have shown that endogenous GLP-1 in humans with and without diabetes contributes to beta-cell secretory response to oral glucose or mixed meal ingestion. Further, rerouted GI anatomy after GB (7, 13, 15–18) or SG (12) increases prandial GLP-1-stimulated insulin secretory response in these cohorts. However, the glucose-dependency of insulinotropic effect of GLP-1 in the fed state in humans with or without bariatric surgery is largely unknown.

The present study was designed to examine the role of endogenous GLP-1 on glucose metabolism and islet-cell function before and after protein ingestion, where glycemic concentrations are not changing from basal values, and determine whether GB exaggerates beta-cell or glycemic effects of GLP-1 during protein intake, as well as to evaluate the differences between GB and SG on these outcomes. Whey protein was used given its potency on the insulin response compared to other protein-containing compounds (24, 25). Tight regulation of pancreatic beta-cells that monitor and respond to ingested glucose and non-glucose nutrients is essential in normal control of glucose homeostasis. In healthy humans, insulin secretion increases in a dose-dependent manner in response to enteral (25, 26) or parenteral amino acid administration (27) while glucose concentration declines or remains at basal values, indicating that amino acids can directly stimulate insulin secretion. Although, a previous observation (28) that oral ingestion versus intravenous infusion of amino acid mixture elicits a much larger insulin secretion at matched circulatory levels of amino acids indicates that gut-derived factors also play a role in beta-cell response after protein ingestion. An incretin role for GLP-1, however, was dismissed by this report since plasma concentration of GLP-1 remained unchanged after amino acid ingestion (28). Thus, we examined the contribution of GLP-1 to insulinotropic effect of oral protein load by using intravenous infusion of GLP-1R antagonist. Blocking GLP-1R resulted in a small but significant increase in glucose concentrations during fasting and fed conditions by ∼5% and ∼10%, respectively, in all 3 groups (Fig.1a inset). The glycemic enhancement in the first 60 min of protein intake, however, was much more prominent in GB and SG than controls (Fig.1a).

The glycemic effect of Ex-9 in fasting state, when there is no nutrient stimulation of GLP-1 secretion, was associated with reduction in insulin secretion in non-operated controls, but not in GB or SG. In the context of recent reports that GLP-1 is produced in the alpha-cell (29), ∼15% insulin reducing effect of Ex-9 infusion in fasting state is consistent with a paracrine, alpha-cell to beta-cell, model of communication regulating insulin secretion similar to what has been previously reported (8, 9). Alpha-cell GLP-1 production is upregulated under metabolic stress, such as obesity and diabetes (30). Consistent with our previous report (7), in the current experiment, the relative effect of GLP-1R blockade on fasting insulin secretion was larger in controls than matched GB or SG subjects. These findings, hint towards a potential downregulation of alpha-cell GLP-1 production by bariatric surgery, but this hypothesis merits further investigation.

In contrast to fasting state, prandial glycemic effect of GLP-1R blockade was associated with a similar reduction in beta-cell sensitivity to glucose in the early phase of protein ingestion (Fig.2a), consistent with an incretin role for GLP-1 during protein ingestion in surgical and non-surgical obese subjects alike. Nonetheless, the overall beta-cell insulin secretory response to protein intake (AUC ISR_3h_) was reduced by ∼30% in GB and SG and increased by ∼20% in controls during Ex-9 infusion despite a similar increase in glucose concentrations (Fig.1b), indicating a larger insulinotropic effect of GLP-1 in GB and SG than controls in the fed state. However, aligned with prior meal studies (7), we did not find any association between the size of GLP-1-stimulated ISR and plasma levels of GLP-1 after protein intake. This observation, the dissociation of plasma GLP-1 concentrations and the insulinotropic effect of GLP-1 where glycemic concentrations are maintained at baseline, indicates that activation of GLP-1R cannot be explained by changes in plasma concentrations of the peptide or glucose.

Recently the endocrine function of GLP-1 in stimulating prandial insulin secretion, particularly after bariatric surgery, has been challenged. In a rodent model of obesity, glycemic improvement of sleeve gastrectomy is mediated by pancreatic rather than intestinally secreted GLP-1(19). Thus, our findings in the context of recent reports (9, 19, 22) raise the possibility that beta-cell effect of GLP-1 to increasing amino acids in euglycemic condition, is mediated by the paracrine action of pancreatic produced peptide rather than intestinally secreted GLP-1 which makes up majority of circulatory concentrations of this peptide.

Furthermore, in our experiment, beyond a greater glucose-independent insulinotropic effect of GLP-1 in surgical versus non-surgical subjects, Ex-9 infusion also increased whole body insulin action (mainly reflecting skeletal muscles) after bariatric surgery (Fig.5b). Therefore, disposition index, a product of insulin secretion and insulin action remained unaffected by administration of GLP-1R antagonist among 3 groups (Table 2). It has previously been shown that ingestion of oral glucose compared to intravenous glucose administration in obese subjects blunts insulin action and diminishes insulin efficacy in suppressing lipolysis despite a higher insulin secretory response (31) but the gut factor responsible for reduced prandial insulin action has not been identified. In our study, insulin action after protein ingestion is diminished by endogenous GLP-1 after GB and SG.

In mice, acute infusion of Ex-9 into the lateral ventricle of the brain has been shown to increase whole body insulin sensitivity (M/I) by 300% and reduce prandial insulin secretory response by 60% (23). Improved peripheral insulin sensitivity by Ex-9 in these experiments was eliminated by muscle denervation (23), suggesting that GLP-1 effect on insulin action is mediated by neural input to muscles. While the translational significance of these findings in humans is difficult to establish, it is also well recognized that GLP-1 signal is detected by visceral afferent nerves in hepatoportal (32) or directly in the central nervous system (23, 33), Therefore, it is plausible that higher intestinally produced GLP-1 secretion due to faster nutrient flux after bariatric surgery can provoke the neural-mediated pancreatic and extra-pancreatic GLP-1 actions in this population. While this hypothesis require further investigation, an exaggerated extra-pancreatic effect of GLP-1 on insulin action in GB-treated subjects could explain the previously reported discord in Ex-9 effect on prandial insulin and glucose response, where a significant reduction in insulin secretion by GLP-1R blockade in GB-treated subjects is not reciprocated by an increase in plasma glucose concentration (16–18).

In addition to the differences in insulin secretion and insulin action among surgical and non-surgical subjects, protein ingestion was associated with a marked stimulation of EGP compared to controls in whom no change in EGP was observed, possibly due to increased gluconeogenesis (34). Further, blocking GLP-1R increased prandial EGP in GB subjects but not in SG or CN (Fig.4b). In healthy individuals, exogenous GLP-1 infusion during a euglycemic (35) or hyperglycemic clamp (36, 37) in the fasting state diminishes EGP, independent of plasma insulin and glucagon concentrations, suggesting a direct effect of GLP-1 on liver glucose metabolism. Our study design cannot distinguish between a direct versus indirect contribution of endogenous GLP-1 to hepatic glucose output given the differences in plasma insulin and glucagon concentrations among the groups and between the studies performed with and without Ex-9. Nonetheless, the absolute differences in prandial glucagon-to-insulin ratio between the two studies was minimal in controls and, while it was increased in surgical groups, it was almost identical between the GB- and SG-treated subjects in our experiment (Supplementary.Figure2). Yet, blocking GLP-1R increased EGP in GB and not in SG, suggesting that either hepatic sensitivity to insulin and glucagon is altered after GB compared to SG or that the GLP-1 effect on EGP in GB is independent of hormonal factors.

Finally, blocking GLP-1R diminished the effect of insulin to suppress plasma FFA in the latter phase of protein absorption in GB subjects (Fig.5c), similar to the effect of Ex-9 infusion on EGP after GB (Fig.4b). It is unclear whether prandial FFA is directly or indirectly affected by Ex-9 infusion, although, based on previous reports, neither lipolysis nor FFA concentrations are changed by *exogenous* GLP-1 or GLP-1R agonist administration during hyperglycemic clamp (37) or oral glucose challenge (38), respectively. Nonetheless, reduction in FFA flux to the liver by *endogenous* GLP-1 observed in our experiments, can also contribute to EGP lowering effect of this peptide, as previously suggested in non-surgical individuals (39).

There are several limitations to this study. We used a cross-sectional rather than longitudinal design which imposes limitations on the effect of weight loss surgery on the outcomes of interest. Nonetheless, using this method, we were able to compare the outcomes in bariatric surgical subjects when they were completely adapted to the metabolic effects of these procedures beyond the first 2 years. GB-treated subjects had a larger weight loss than SG subjects, mainly due to weight loss in the first 6-12 months of their surgery, but the current BMI was similar among the groups and the participant’s body weight was stable for 3 months prior to study. We did not measure the plasma amino acid concentration; however, it can be assumed, based on previous studies (40) there is an earlier and higher peak amino acid concentration after GB compared to SG and in SG subjects versus controls.

In conclusion, our novel observation demonstrates that GLP-1 plays an important role in the stimulatory effect of oral protein ingestion on insulin secretion in humans and that this action is independent of the plasma glucose concentration. Further, rerouted GI anatomy after gastric bypass or sleeve gastrectomy not only augments the stimulatory effect of GLP-1 in insulin secretion but also provokes extra-pancreatic action of GLP-1. As demonstrated by the present results and consistent with new evidence from clinical and preclinical studies, there are important effects of GLP-1 signaling on metabolic homeostasis that occur independently of the plasma levels of peptide and make a case for paracrine action of GLP-1 produced by islet-cells or neural-mediated action of this peptide produced by intestinal L-cells or central nervous system. Our results lay the foundation for future mechanistic studies to examine the relevance of the hormonal and non-hormonal GLP-1 signals on multi-targeted treatment approach utilizing surgical, medical, and nutritional interventions or combinations thereof for treatment of diabetes and obesity.

## METHODS

### Sex as a biological variable

Both men and women were recruited in this study. However, in keeping with the national estimate of higher ratio of females to males after bariatric surgery (105), we aimed for a 60-70% ratio of females to males in our cohort.

### Subjects (Table1)

Ten non-diabetic individuals with previous history of GB and 9 BMI- and age-matched subjects with SG and 7 healthy non-operated CN were consecutively recruited based on their response to our enrollment effort. None of the participants had diabetes or renal dysfunction or liver disorder. The control subjects had no personal or family history of diabetes and had a normal oral glucose tolerance test. Subjects were weight stable for at least 3 months prior to enrollment.

### Peptides

Synthetic exendin-(9–39) (CS Bio, Menlo Park, California) was greater than 95% pure, sterile, and free of pyrogens. Lyophilized peptide was prepared in 0.25% human serum albumin on the day of study. The use of synthetic exendin-(9–39) is approved under the U.S. Food and Drug Administration Investigational New Drug 123,774.

### Experimental procedures

Subjects were instructed to eat a weight-maintaining diet containing 150-200 grams of carbohydrates per day and not to engage in vigorous physical activity for 3 days prior to each study visit. Studies were performed at the Bartter Clinical Research Unit at Audie Murphy VA Hospital in the morning after an overnight fast. Body composition was assessed using dual-energy X-ray absorptiometry, and waist circumference was measured. Intravenous catheters were placed in each forearm for the blood withdrawal and the infusion of study drugs; the arm used for blood sampling was continuously warmed using a heating pad to arterialize the venous blood. Blood samples were drawn from -130 to 180 minutes; the plasma was separated within 60 minutes for storage at -80°C until assay.

At -120 minutes, a primed-continuous infusion of [6,6-^2^H_2_] glucose (28 µmol/kg prime and 0.28 µmol/kg/min constant) was initiated and continued for the duration of the study as previously described (7). At -60 minutes, subjects either received a primed continuous infusion of Ex-9 (7,500 pmol/kg prime and 750 pmol/kg/min constant) or saline for the remainder of the study (7). At time 0 min, 50 g whey protein mixed with 1 g of acetaminophen was consumed orally within 10 min. The order of the studies was performed in random fashion.

### Assays

Blood samples were collected in EDTA tubes for measurement of insulin, acetaminophen, glucose and in aprotinin/heparin/EDTA for assay of C-peptide, glucagon, GLP-1, and GIP (41). Plasma glucose was determined using Analox GM9 Glucose Analyzer (Analox Instruments, Stourbridge, UK). Insulin (DIAsource, Neuve, Belgium), C-peptide and glucagon (Millipore, Billerica, MA) were measured with commercial radioimmunoassay kits. The Millipore glucagon RIA kit has a cross-reactivity of <2% with oxyntomodulin and glicentin with a sensitivity of ∼10 pmol/l (42). GIP was measured using commercial Multiplex ELISA (Millipore, Billerica, MS), and GLP-1 using ELISA (Mercodia, Uppsala, Sweden) according to the manufacturers’ specifications. Tracer enrichment was measured by GC-MS (5975, Agilent, Santa Clara, CA) as previously described (7, 43, 44) utilizing the same derivatization method used for glucose tracers and monitoring peak of mass 200-202. Acetaminophen was measured by GC–MS using acetaminophen (acetyl-^13^C_2_,^15^N) as internal standard (Cambridge Isotope Laboratories, Boston, USA) free fatty acid (FFA) was determined by calorimetric assay (Wako Chemicals, Richmond, VA, USA).

### Calculations

Fasting plasma glucose and hormone concentrations represent the average of 2 samples drawn before -120 min, and the pre-meal values represent the average of 2 samples drawn before the test meal. Insulin secretion rates (ISRs) were calculated from C-peptide concentrations using deconvolution with population estimates of plasma C-peptide (45). Beta-cell glucose sensitivity was calculated as the slope of ISR and blood glucose concentration for the first part of protein absorption, as ISR rose to peak value. Beta-cell sensitivity to GLP-1 was measured as the slope of each subject’s plot of ISR (from premeal to peak values) versus corresponding plasma concentration of GLP-1(46).

Rates of total glucose appearance (*Ra*), reflecting endogenous glucose production (*EGP*), and total glucose disappearance (*Rd*) were derived from plasma [6,6-^2^H_2_]glucose enrichments as previously described using the Steele equation (7, 43). Metabolic clearance of glucose (*MCG*) was measured as *Rd*/plasma glucose (31, 44).

Using the trapezoidal rule, the prandial incremental area under the concentration curve (AUC) of islet-cell and gut hormones, as well as glucose fluxes, was calculated from 0-60 and 0-180 minutes to examine the early and total responses, respectively, given the altered prandial response pattern after bariatric surgery. Pre- and post-prandial insulin sensitivity were calculated as the ratio of premeal *MCG*/insulin and the prandial total AUC of the *MCG*/insulin, respectively (31).

Insulin extraction and clearance rates were calculated as previously described (45). Disposition index was calculated as the product of total AUC ISR and *MCG*/insulin during the 3 hours after oral glucose ingestion (31, 44). Antilipolytic effect of insulin was measured as FFA per unit of insulin, i.e., the ratio of premeal free fatty acid (FFA)/insulin and the prandial total AUC of the FFA/insulin, given the linear relationship between the two parameters within insulin range in our experiments(46).

## STATISTICAL ANALYSIS

Data are presented as mean ± SEM. The parameters of interest at baseline and the relative changes in the outcomes from saline to Ex-9 study were compared using ANOVA or Chi-square. The effect of administration of GLP-1R antagonist and the group effect (GB, SG, and CN), as well as their interaction on experimental outcomes, were analyzed using repeated measured ANOVA with post-hoc (Tukey’s) comparisons among the groups. Association among parameters were performed using Spearman correlation. Statistical analyses were performed using SPSS 28 (SPSS Inc., Chicago, IL). The STROBE cross sectional reporting guidelines were used (47).

## STUDY APPROVAL

The Institutional Review Board of the University of Texas Health at San Antonio approved the protocol (HSC20180070H) and all subjects provided written informed consent before participation.

## AUTHOR CONTRIBUTIONS

M.R. performed data analysis and wrote the original draft of the manuscript. A.G. undertook mathematical modeling and wrote the original draft of the manuscript. H.H conducted the studies. S.P. and F.C. assisted with mathematical modeling. R.P. assisted in the recruitment of participants. R.D. reviewed/edited the manuscript. M.S. designed the study, oversaw its conduct, performed data analysis, and reviewed/edited the manuscript. Dr. Marzieh Salehi is the guarantor of this work and, as such, had full access to all the data in the study and takes responsibility for the integrity of the data and the accuracy of the data analysis. The order of co-first authors was determined by the time that each joined the project.

## Data Availability

All data produced in the present study are available upon reasonable request to the authors.

## ACKNOWLEDGEMENTS

We thank Andrea Hansis-Diarte, Nancy Yegge, and John Adams from the Department of Medicine of University of Texas Health at San Antonio for their technical support and nursing staff as well as nutritionist from Bartter Research Unit, Audie Murphy Hospital, STVHCS, for their expert technical assistance. We owe a great debt to our research participants.

## Supplementary Figure Legends

**Supplementary Figure1**. (a) Absolute levels of glucagon-to-insulin ratio after oral protein load with (dashed line) and without (solid line) GLP-1R blockade (Ex-9), and (b) difference in glucagon-to-insulin ratio between the two studies of saline and Ex-9, in subjects who underwent gastric bypass (black line) or sleeve gastrectomy (red line) and non-operated controls (blue line).

**Supplementary Figure2.** Plasma concentrations of acetaminophen during protein ingestion with (dashed line) and without (solid line) intravenous infusion of exendin-(9–39) (Ex-9), GLP-1R antagonist, in subjects who underwent gastric bypass (black line) or sleeve gastrectomy (red line) and non-operated controls (blue line).

