## Supplementary figures and images for "GLP-1 enhances beta-cell response to protein ingestion independent of glycemia and bariatric surgery amplifies it"

### MEDRXIV 2024 Supplementary Figure 1.gif

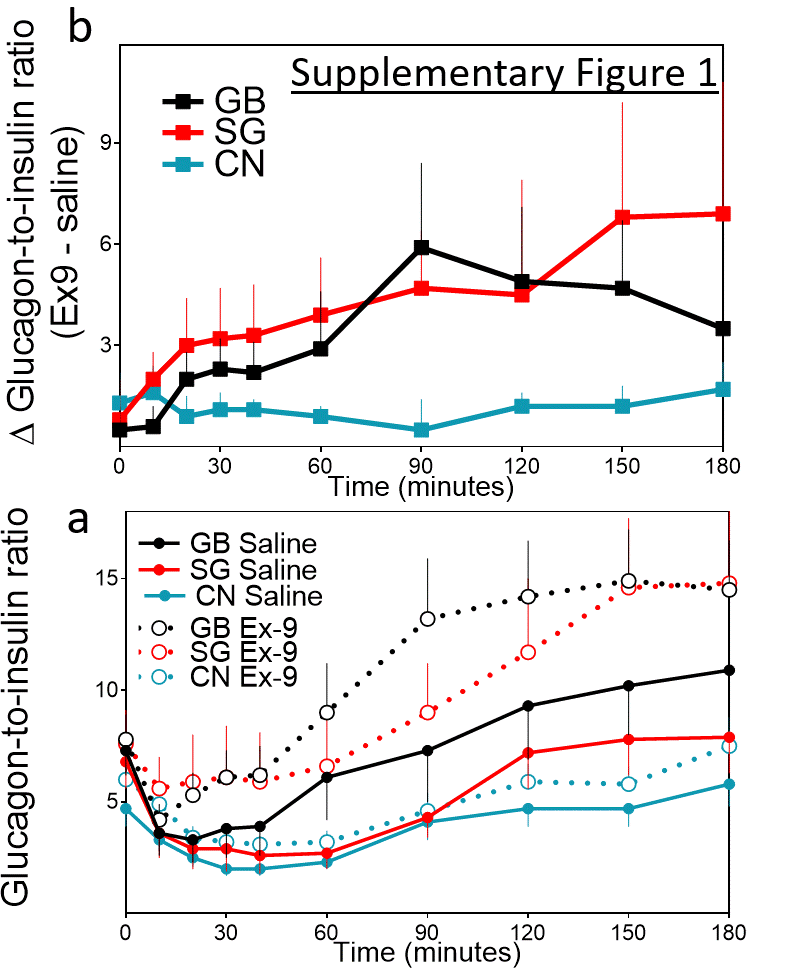

### MEDRXIV 2024 Supplementary Figure 2.gif

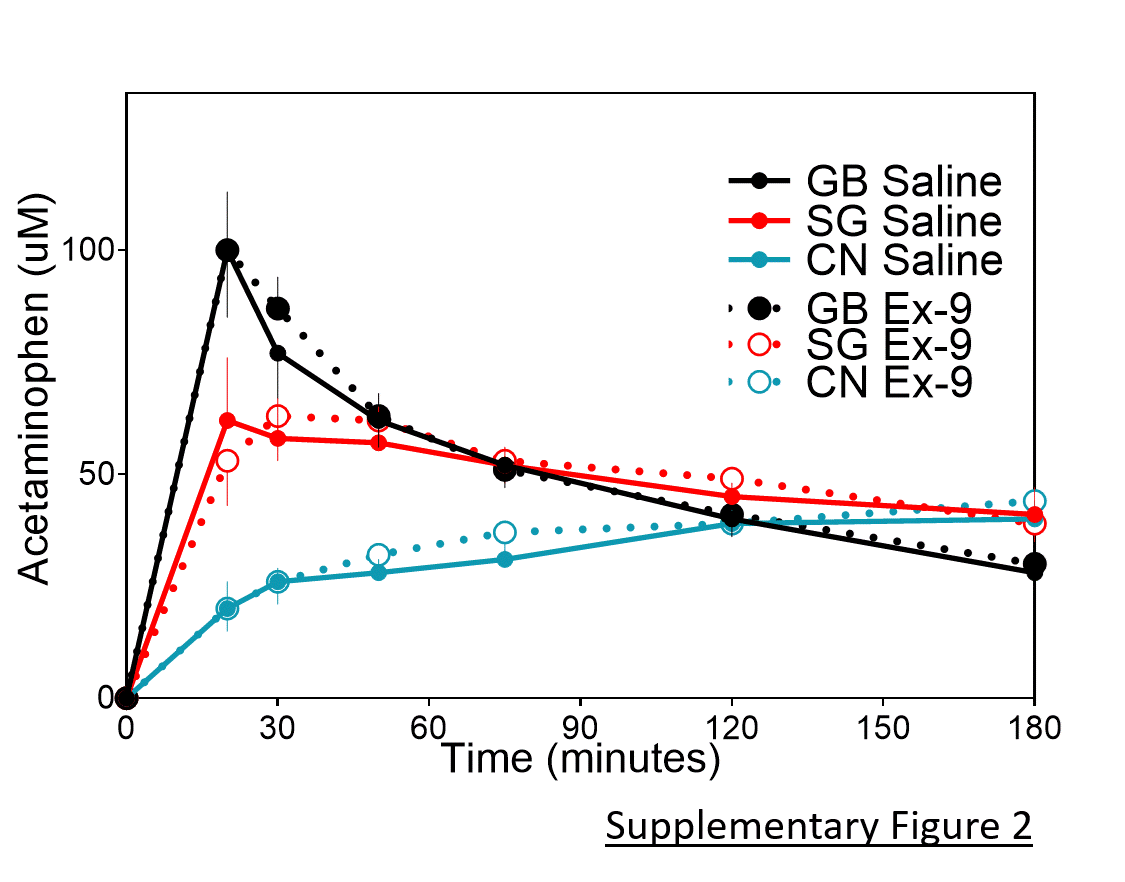
